# Zero-Shot Multimodal Question Answering for Assessment of Medical Student OSCE Physical Exam Videos

**DOI:** 10.1101/2024.06.05.24308467

**Authors:** Michael J. Holcomb, Shinyoung Kang, Ameer Shakur, Sol Vedovato, David Hein, Thomas O. Dalton, Krystle K. Campbell, Daniel J. Scott, Gaudenz Danuser, Andrew R. Jamieson

## Abstract

The Objective Structured Clinical Examination (OSCE) is a critical component of medical education whereby the data gathering, clinical reasoning, physical examination, diagnostic and planning capabilities of medical students are assessed in a simulated outpatient clinical setting with standardized patient actors (SPs) playing the role of patients with a predetermined diagnosis, or case. This study is the first to explore the zero-shot automation of physical exam grading in OSCEs by applying multimodal question answering techniques to the analysis of audiovisual recordings of simulated medical student encounters. Employing a combination of large multimodal models (LLaVA-1.6 7B,13B,34B, GPT-4V, and GPT-4o), automatic speech recognition (Whisper v3), and large language models (LLMs), we assess the feasibility of applying these component systems to the domain of student evaluation without any retraining. Our approach converts video content into textual representations, encompassing the transcripts of the audio component and structured descriptions of selected video frames generated by the multimodal model. These representations, referred to as “exam stories,” are then used as context for an abstractive question-answering problem via an LLM. A collection of 191 audiovisual recordings of medical student encounters with an SP for a single OSCE case was used as a test bed for exploring relevant features of successful exams. During this case, the students should have performed three physical exams: 1) mouth exam, 2) ear exam, and 3) nose exam. These examinations were each scored by two trained, non-faculty standardized patient evaluators (SPE) using the audiovisual recordings—an experienced, non-faculty SPE adjudicated disagreements. The percentage agreement between the described methods and the SPEs’ determination of exam occurrence as measured by percentage agreement varied from 26% to 83%. The audio-only methods, which relied exclusively on the transcript for exam recognition, performed uniformly higher by this measure compared to both the image-only methods and the combined methods across differing model sizes. The outperformance of the transcript-only model was strongly linked to the presence of key phrases where the student-physician would “signpost” the progression of the physical exam for the standardized patient, either alerting when they were about to begin an examination or giving the patient instructions. Multimodal models offer tremendous opportunity for improving the workflow of the physical examinations’ evaluation, for example by saving time and guiding focus for better assessment. While these models offer the promise of unlocking audiovisual data for downstream analysis with natural language processing methods, our findings reveal a gap between the off-the-shelf AI capabilities of many available models and the nuanced requirements of clinical practice, highlighting a need for further development and enhanced evaluation protocols in this area. We are actively pursuing a variety of approaches to realize this vision.

## 1 Introduction

An Objective Structured Clinical Examination (OSCE) is a critical component of Medical Education whereby the data gathering, clinical reasoning, physical examination, diagnostic and planning capabilities of medical students are assessed in a simulated outpatient clinical setting with standardized patient actors (SPs) playing the role of patients with a predetermined diagnosis, or case. After soliciting a chief complaint from the patient, a student should collect information about the simulated patient’s present illness and their medical, family, and social histories, perform pertinent physical exams, and offer a diagnosis and an appropriate treatment plan.

These examinations provide a rich set of performance data for these students, ensuring they are competent and well-trained before providing actual patient care. However, due to their unstructured nature, these examinations are difficult to grade. Currently, most administrators of these exams rely on highly specialized, experienced teams to manually assess each student’s performance, which requires significant time and effort.

One of the most time-intensive elements of this assessment is determining whether students performed all of the relevant physical examinations for the case during their exercise and the degree to which each of these examinations was correctly performed. This work explores a proposed method to interrogate videos with the question, “*Did a student perform an exam?*” using only the audiovisual recording of a student’s exercise.

We frame the problem as a zero-shot, multimodal question-answering task against the unstructured data stream to elicit salient features for answering this question, a precursor to addressing the broader assessment problem. A system of large multimodal models (LMM) and an automatic speech recognition (ASR) model transform the visual and audio information into text, respectively. Subsequently, the proposed technique relies on a large language model (LLM) to query the extracted text information and generate an answer with supporting evidence, depicted in Figure 1 as an abstractive question-answering task.

**Figure 1:**
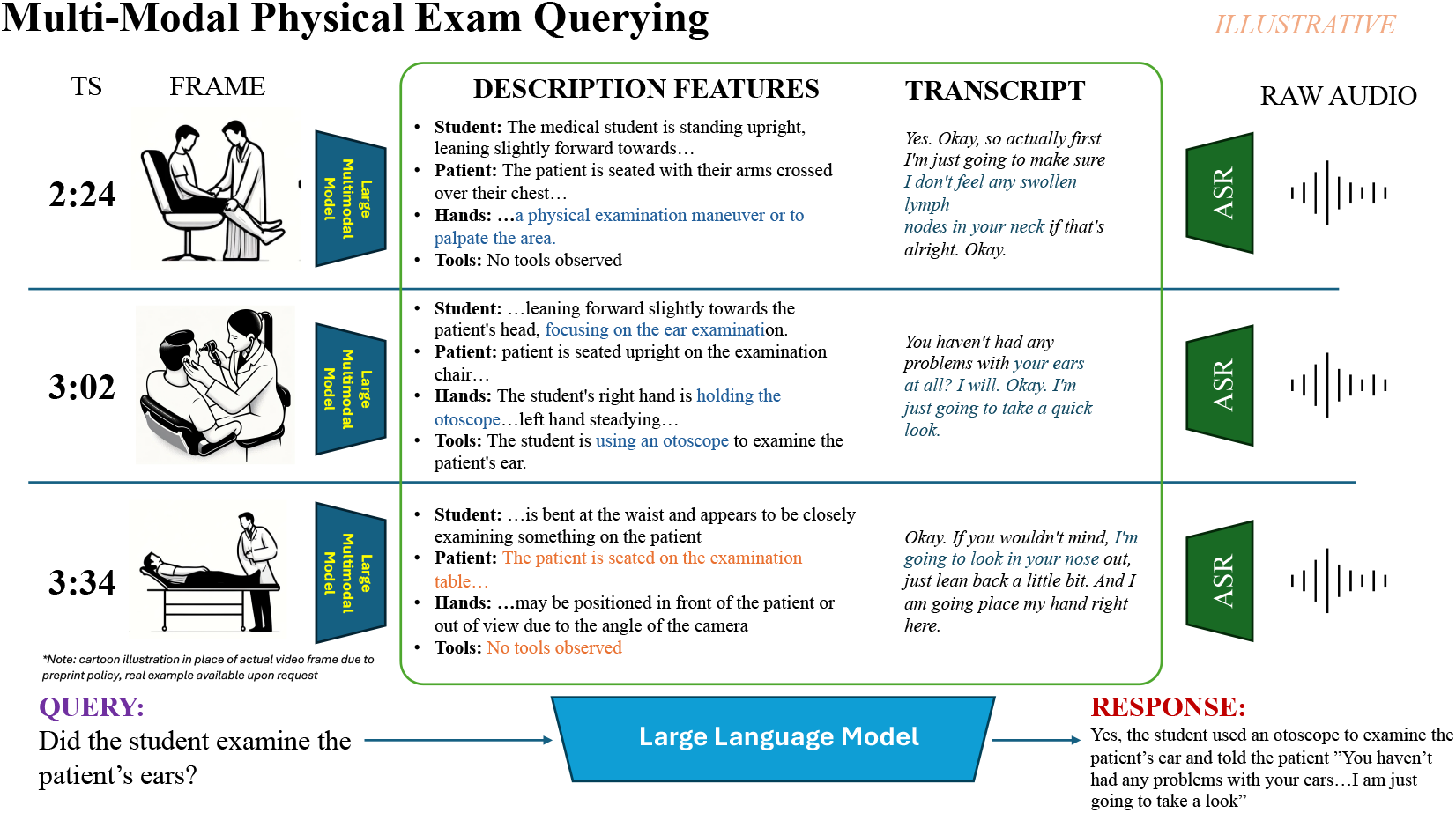
System Illustration - Construction of the “Exam Story”

## 2 Prior Work – Simulation Assessment

Given the resources required for grading these assessments, there have been several efforts to evaluate automating aspects of OSCE administration (Salt et al., 2019). To date, most success has been found in developing systems that grade post-encounter notes, the free-text responses that students write after the simulated encounter. Recent attempts have focused on using supervised learning (Sarker et al., 2019; Yim et al., 2019) that utilize previously scored triples of student responses, rubrics, and scoring data to train binary classifiers or semi-supervised approaches that use clinical entity recognition pipelines and curated acceptable answer lists to test for the presence of key phrases (Bond et al., 2023).

Concerning transcripts of simulated exams, Mistica (2008) explored the use of discourse analytics features in the automated scoring of two selected cases. Fareez (2022) published a collection of simulated patient encounter transcripts for respiratory cases to facilitate ASR methods specific to this domain.

Comparatively, less work has been done on analyzing audiovisual recordings of simulated patient exams. Vedovato et al. (2024) proposed a supervised learning approach for detecting the performance of ear exams from video frames, while Meinich-Bache (2020) demonstrated the development of an object detection system in the setting of neonatal resuscitation simulations that could support downstream assessment and analysis.

## 3 Experimental Setup

### 3.1 Dataset

A collection of 191 audiovisual recordings of medical student encounters with an SP for a single OSCE case was used as a test bed for exploring relevant features of successful exams. We reserved four of the recordings for code and prompt development (the “development set”), leaving the remaining 187 as an evaluation set (the “test” set).

During this case, the students should have performed three physical exams: 1) mouth exam, 2) ear exam, and 3) nose exam. These examinations were each scored by two trained, non-faculty standardized patient evaluators (SPE) using the audiovisual recordings—an experienced, non-faculty SPE adjudicated disagreements. Within our dataset, students appropriately performed the mouth, ear, and nose exams 94%, 75%, and 81% of the time, respectively. For each of the 15-minute recordings, the beginning and ending timestamps of the physical exam portion were manually identified and validated. All the analyses described below were confined to the portion of each recording between these timestamps.

### 3.2 Key Frame Selection

From the visual recordings, a sample of key frames was selected from each video for further analysis with multimodal models. Using a combination of a Detectron2 (Wu et al., 2019) model fine-tuned on hand detection and a pre-trained Detectron2 model for extracting human poses; several metrics were extracted for each frame at five frames per second, such as various distances between the two participants’ body parts (face, neck, hands, etc.).

These frame-level metrics were then clustered using a temporal k-means clustering method, ensuring that video segments were close in feature space and temporally. The key frames were selected by choosing the frame corresponding to the median distance values from the cluster with the shortest ear-to-hand distance. More details on this pipeline can be found in the Appendix.

### 3.3 Transcription

The audio tracks of the recordings were transcribed using OpenAI’s Whisper-large-v3 (Radford et al., 2023), an ASR model. The transcription pipeline was implemented using HuggingFace’s transformers library (Wolf et al., 2020). FlashAttention-2 (Dao et al., 2022) and a batch size of 16 were utilized to accelerate the ASR inference. The timestamped utterances within the previously identified physical exam window of the exercise were used without any further human annotation or correction.

## 4 Methodology

To explore zero-shot comprehension of audiovisual recordings through natural language processing, our approach converts video content into textual representations, encompassing the transcripts of the audio component and structured descriptions of selected video frames. These representations, referred to as “exam stories,” are then used as context for an abstractive question-answering problem. This method relies on the capabilities of LLMs to respond to complex queries over extended contexts. This system is illustrated in Figure 1.

### 4.1 Image Frame Descriptions

For the generation of image frame descriptions, we employed the LLaVA family (Liu et al., 2023) of large multimodal models alongside GPT-4 with Vision and GPT-4o (OpenAI et al., 2023). These models were used to extract structured fields covering the essential elements of each frame, such as the postures of the student and patient, the student’s hand movements, and the presence of any detected tools to be analyzed by downstream natural language processing algorithms. The prompts used and an example frame description are included in the Appendix.

### 4.2 Assessment as Multimodal Question

Our methodology implements evaluation as a multimodal question-answering task in assessing student physical exam performances. We explore using three different constructions of contexts, or “exam stories,” namely image-based, transcript-based, and combined (integrating both image descriptions and transcripts). This stratified approach allows for a comparative analysis of the relative effectiveness of visual and audio features in supporting multimodal action recognition as a textual task.

Each construction of the exam story is analyzed through structured prompts that elicit binary responses (“yes” or “no”), extractive supporting evidence, and the justification of the grading decision. This response structure aids in the post-hoc critique of the assessment determination. Provided in the Appendix are example exam stories and model responses, illustrating the application of our multimodal assessment framework to the grading of student video recordings.

To aid in comparability, the video frame descriptions of each LMM were paired for the final question-answering step with its corresponding LLM backbone (e.g., LLaVA-1.6 Mistral 7B frame descriptions were provided to the Mistral 7B Instruct for question-answering; LLaVA-1.6 Vicuna 13B to Vicuna 13B; LLaVA-1.6 Nous Hermes Yi 34B to Nous Hermes Yi 34B; GPT 4 with Vision to GPT 4 Turbo; GPT 4o itself).

## 5 Results

As shown in **Table 1**, the agreement between the described methods and the SPEs’ determination of exam occurrence as measured by percentage agreement varied from 26% to 83%. The audio-only methods, which relied exclusively on the transcript for exam recognition, performed uniformly higher by this measure compared to both the image-only methods and the combined methods across differing model sizes.

**Table 1:** Percentage agreement with SPEs.

| Model Family | Image | Audio | Combined |
| --- | --- | --- | --- |
| LLaVA/Mistral 7B | 25.5% | 75.2% | 35.7% |
| LLaVA/Vicuna 13B | 36.5% | 80.2% | 65.8% |
| LLaVA/NS Yi 34B | **52.4%** | 78.8% | 76.3% |
| GPT 4 w/ Vision | 34.6% | 82.7% | **80.2%** |
| GPT 4o | 40.6% | **83.2%** | 77.0% |

**Figure 2** depicts a breakdown by exam type of the SPE agreement as measured by Cohen’s Kappa. Most model configurations had the highest agreement on the assessment of the ear exam and the lowest on the nose exam.

**Figure 2:**
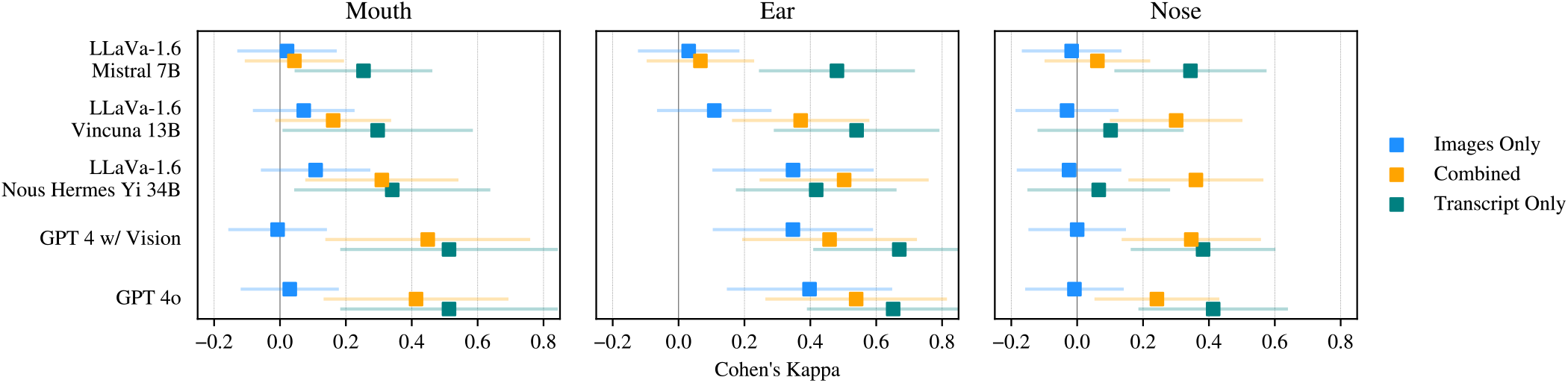
Cohen’s Kappa and 95% confidence intervals by exam type for each model/modality combination.

**Figure 3:**
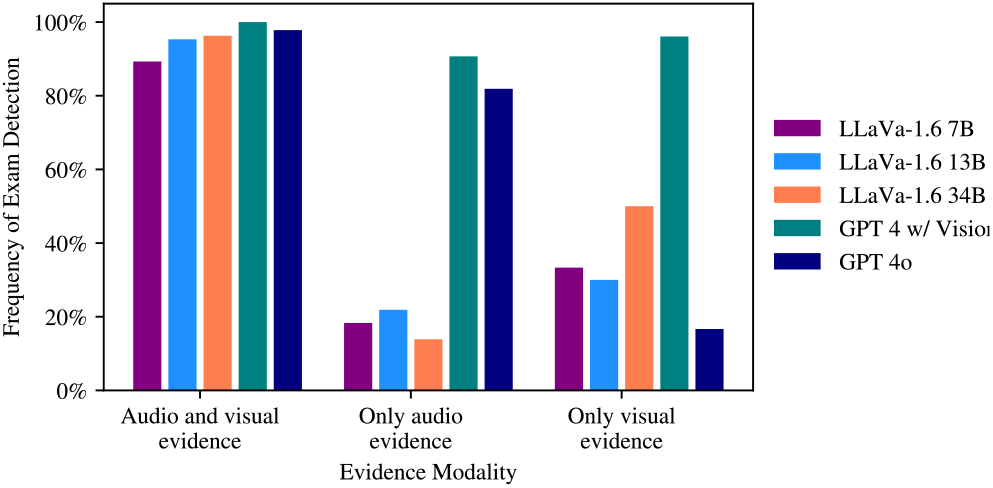
Exam Detection Rates for Combined Models by Evidence Type

Notably, the combined input models outperformed the transcript-only versions in specific settings, such as for the LLaVA/Vicuna 13B nose exam and LLaVA/Nous Hermes Yi 34B ear and nose exams.

## 6 Discussion

### 6.1 Modality Reliance

To understand the relative outperformance of the transcript-only configurations, the reliance on different evidence modalities was analyzed for the models with combined inputs. In the prompts of the combined input configurations, two fields, “visual evidence” and “audio evidence,” were solicited in the response, prompting the models to extract any modality-specific evidence supporting the occurrence of the queried activity; otherwise, the LLM was to report “no visual/audio evidence.” This permitted a high-level analysis to measure how often the model responded in the affirmative for exam occurrence when 1) both input streams contained supporting evidence, 2) only the audio stream had supporting evidence, or 3) only the vision stream supported the exam occurring.

All models predicted the occurrence of an exam nearly every time evidence from both modalities was extracted. However, the GPT 4 with vision model was the only one to frequently predict an exam occurrence when only one modality offered supporting evidence (audio or image only). The other models infrequently predicted an exam occurrence when only the audio stream offered evidence.

### 6.2 Image Description Errors

A review of the image-only model outputs revealed systemic shortcomings in the ability of the LMMs to correctly identify the presence of tools, location of hands, or the correct location of tool placement. All models rarely identified the tools used in the nose exam which partially explains the lower SPE agreement for this exam. In other instances, while the primary tool may have been correctly identified, its usage was attributed to the incorrect exam or action. These errors most likely arise from the over-to-shoulder perspective of the videos, and the limited amount of clinical data in the training datasets. OpenAI (2023) cautions explicitly against using its model in the medical domain and with bird’s-eye view camera angles. A detailed breakdown of the image frame description errors can be found in the Appendix.

### 6.3 Student-Physician Signaling

The outperformance of the transcript-only model was strongly linked to the presence of key phrases where the student-physician would “signpost” the progression of the physical exam for the standardized patient, either alerting when they were about to begin an examination or giving the patient instructions. SPEs awarded students credit for the corresponding exam every time one of the following phrases could be detected in the transcript: “Open your mouth…” for mouth exams; “…take a look at your ear.” for ear exams; and “…. take a look in your nose.” for nose exams.

While these signposts were highly correlated with SPE grading, the phrases themselves provide no further information about how well the student performed the physical exam, if at all. As such, the simple measure of SPE agreement alone is insufficient evidence that a system achieves the desired assessment performance goals in this setting.

## 7 Conclusion

This study explores the potential of automating OSCE grading through a zero-shot multimodal question-answering framework. Despite the promising correlations with human expert evaluations in specific contexts, this research highlights a significant limitation of general purpose LMMs: their inadequacy in accurately interpreting over-the-shoulder videos within clinical settings. This challenge is emblematic of the broader difficulty in automating nuanced tasks such as medical assessments, where the subtlety of clinical activities remains outside the domain of the examined models without adaptation.

Our findings underscore the necessity for further development and refinement of LMMs, specifically tailored to the clinical domain to overcome these obstacles and the development of more robust protocols for system evaluation. The study’s insights into the differential effectiveness of audio versus combined modality inputs offer a valuable starting point for such advancements and highlight the dangers of high-correlation features such as signposting that on their own do not evidence the successful performance of an exam.

### Limitations

The dataset used for this study was confined to the recordings of a single site administration of OSCEs for medical students over one semester for one case. These recordings were collected from a limited number of rooms and camera configurations over seven non-consecutive days. The limited size of the dataset may also limit the generalizability of the findings as well as the limited number of physical exam types evaluated; however, exams performed and assessed mirrored the national consensus of physical exam standards by the National Board of Medical Examiners’ United States Medical Licensing Examination (USMLE) (United States Medical Licensing Examination, n.d.).

The assessment process evaluated here is a collection of specific prompts with structured outputs; the performance of such a system is expected to be sensitive to different prompting strategies, especially those with more straightforward or complex structured responses. Furthermore, the complexity of the current structured responses resulted in numerous parsing failures; the evaluations here do not contemplate possible improvements due to retrying generations with parsing errors. The unsupervised key frame selection method may not always select the best individual frames within a longer video. This may have contributed to some disagreements with SPEs irrespective of the underlying LMMs employed.

Only four LMMs from two different model families were evaluated which may have low correlation with LMMs of other families. Additionally, all evaluations were done in a zero-shot fashion which provides no supporting data for how model behavior may change with in-context learning or fine-tuning. The exam story inputs to the LLMs often exceeded 4,000 tokens, necessitating the use of more extended context models or RoPE scaling; differences in model adaptation to extended contexts may have contributed to observed differences in model performance.

The primary evaluation method was measuring agreement with SPEs which limits any inferences as to the quality of the exam performance or the accuracy of the model scoring rationales. While an item-level error assessment was performed on the image-only models, showing a fundamental weakness with tool identification, these error mixes may not necessarily translate across the other modalities or exam types. While up to three non-faculty SPEs graded each of these exams, no comparisons were made between model predictions and physician assessments of exam performance.

## Data Availability

Raw OSCE audio-video recording data of the students (FERPA protected) are not publicly available. Privacy sensitive analytical outputs may be made available upon request to authors.

## A Preprocessing Algorithms

### Doctor Patient Body Separation

During our OSCE set-up, patients were already present in the examination room when doctors entered. To distinguish between the two, we assigned the first human data to the patient’s body and labeled the new human body that would appear later in the video as the doctor’s body. On the assumption that there could only be a maximum of two humans in the frame, each new human body information obtained from later frames was matched to either the patient’s or doctor’s body according to the dot product of minimum distance for each body part.

### Key Frame Selection

We analyzed the physical exam at a higher sampling rate of five frames per second to study the dynamics of the doctor’s hands during the examination. We used a pre-trained Detectron2 model to detect human poses and fine-tuned Detectron2 models to detect hands and tools within each frame. The first frame of the doctor and patient was obtained from the previous one-frame-per-second run on the entire video. After that, we differentiated the human body data of the patient and the doctor using the same algorithm used for doctor-patient body separation.

We measured the distance of the closest midpoint of the doctor’s hand to the patient’s nose, ears, and neck (midpoint of the shoulders) throughout the physical exam. We used k-means clustering to group the data into three clusters and identified the cluster with the shortest mean distance between the patient’s ear and the hand. We then marked the timepoint corresponding to the median of the cluster’s ear-to-hand distance as a representative frame of that cluster.

### Cropping

Once we selected the frames for each video, we used the Detectron2 model to identify the minimum bounding box that encloses the doctor’s and patient’s actions in the frame. We then resized this box to 672 × 672 pixels, which served as the input for the vision models. Since the otoscope exam is performed near the patient’s head, if the combined bounding box exceeded 672 pixels in height, we removed the bottom portion of the box. If the image dimensions were smaller than 672 pixels in either height or width, we increased the size of the image by its center bidirectionally, as much as possible. After resizing, we save the images in JPEG format under each video file.

## B Text Generation Specifications

Here we briefly describe the language models and inference setup used to conduct our experiments.

### LMMs – Image Descriptions

- **LLaVA 1.6 7B, 13B, and 34B** (Liu et al., 2023) are a family of large multimodal models that integrate a vision encoder with a large language model to address visual understanding tasks. We adapted the model inference code published by (Liu et al., 2023) to perform inference with bloat16 precision, FlashAttention-2 (Dao et al., 2022), and beam search decoding with a beamwidth of 5. The models were served using a single compute node with 4 H100 GPUs and tensor parallelism across all available GPUs. All other parameters and decoding hyperparameters were left as published.
- **GPT-4 Turbo with Vision and GPT-4o** (OpenAI et al., 2023) are large multimodal models developed by OpenAI that can respond to queries over textual and visual inputs. We queried the hosted “vision-preview” model version in Microsoft Azure’s US West region with all content filters disabled for GPT 4 with Vision and GPT 4o. Model inference requests were made via the chat completion API using the “2024-02-15-preview” and “2024-02-01” API versions for GPT 4 with Vision and GPT 4o, respectively, with a seed of 42 and temperature of 0 to limit, but not prevent, stochasticity of responses. These settings were also used when prompting for the Exam Story Question Answering. All images were passed using the “high” detail setting in a JPEG encoded, base 64 string.

### LLMs – Exam Story Question Answering

- **Mistral 7B Instruct v0.2** (Jiang et al., 2023) is an instruction fine-tuned open weights model from Mistral AI. We used the vLLM (Kwon et al., 2023) inference engine to serve the model with float16 precision and a default context window of 8,192 tokens. The model was served on a V100 GPU with 32GB of VRAM.
- **Vicuna-13B-16k v1.5** (Chiang et al., 2023) is an instruction fine-tuned model based on the Llama 2 architecture with linear RoPE scaling to extend the context window to 16 thousand tokens. We utilized the FastChat (Zheng et al., 2023) platform to serve the model on 2 V100 GPUs.
- **Nous Hermes Yi 34B** (Nous Research, 2023) is an instruction fine-tuned version of Yi 34B (AI et al., 2024) open weights model. We used linear RoPE scaling to extend the maximum context window from of this model from 4,096 tokens to 8,192 tokens. The model was served in float16 precision using a vLLM server on a single node with 4 V100 GPUs.

For each of these models, we used a generation configuration that used a greedy decoding strategy.

**Table 2:** Response parsing failure rates for each input modality and model.

| Model | Audio Only | Image Only | Combined |
| --- | --- | --- | --- |
| Mistral-7B Instruct 0.2 | 2.58% | 0.48% | 0.79% |
| Vicuna-13b-v1.5 16k | 2.74% | 0.00% | 2.22% |
| Nous-Hermes-2 Yi-34B | 1.13% | 0.16% | 4.44% |
| GPT 4 with Vision | 0.00% | 0.00% | 0.00% |
| GPT 4o | 0.00% | 0.00% | 0.00% |

All responses that failed JSON parsing with the specified key values were treated as negative responses. We found that response parsing failures occurred with the following frequencies:

## C Prompts

1. Image Frame Description Prompt:

~~~
<SYSTEM>
You are helpful teaching assistant at a medical school who is helping to assist in the grading of objective structured clinical examinations. You will be provided with frames from a video recording of a medical student interacting with a standardized patient (actor) in an exam room. Your task will be to help describe what is within the frame and provide specific details.
<USER>
{image_data}
TASKS:
1. Describe what can be seen within the video frame and remark on the general quality of view the participants and the action.
2. Grade the quality of the camera angle from (A) excellent to (C) poor; (C) the participants faces and hands are not clearly visible, (B) the participants faces and hands are mostly visible, (A) the participants faces and hands are clearly visible.
3. Medical Student’s Posture: Describe the student’s position and posture relative to the patient
4. Instruments or Tools: Describe any tools or instruments that the student is using. If none, say “No tools observed”; Students may be expected to use any of the following: penlight, otoscope, ophthalmoscope, nasal speculum, or tongue depressor
5. Patient’s Posture: Describe the patient’s position and posture relative to the student and the room
6. Medical Student’s Hand Movements: Describe any hand movements that the student is using within the frame, specifically if they are interacting with the patient. If nothing notable, say “No notable hand movements”
RESPONSE FORMAT:
Please provide a response in the following format as a JSON object with the “description”, “student_posture”, “tools”, “patient_posture”, and “hand_movements” keys.
{{
     “description”: “general description of the video frame and quality”,
      “camera_angle”: “A” | “B” | “C”,
      “student_posture”: “describe the student’s posture and position within the room”,
      “tools”: “describe any tools that are visible and being used within the frame by the student”
      “patient_posture”: “describe patient’s posture and position within the room”,
      “hand_movements”: “describe any hand movements that the student is using within the frame”
~~~

2. Transcript-Only Physical Exam Assessment Prompt

~~~
<SYSTEM>
You are helpful teaching assistant at a medical school who is helping to assist in the grading of objective structured clinical examinations. You will be provided with summary descriptions of frames from a video recording of a medical student interacting with a standardized patient (actor) in an exam room. Your task will be to help ascertain which exams were performed.
<USER>
## EXAM STORY:
{exam_story}
## QUESTION:
During the exam described above with audio transcripts, is the physician performing an exam of the patient’s {exam_type}, yes or no? If so, describe what in the audio transcripts supports this answer in the fields below. If no audio transcript support, say “no audio evidence supports performance of exam”“.
## RESPONSE FORMAT:
Please ONLY provide a response in the following format as a JSON object with the “audio_evidence”, “rationale” and “answer” keys and values as described.
{{
      “audio_evidence”: <summary of transcription evidence supporting presence or absence of exam>,
      “rationale”: <summary of rationale supporting final determination of whether the exam was performed>,
      “answer”: “yes” | “no”
~~~

3. Image-Only Physical Exam Assessment Prompt

~~~
<SYSTEM>
You are helpful teaching assistant at a medical school who is helping to assist in the grading of objective structured clinical examinations. You will be provided with summary descriptions of frames from a video recording of a medical student interacting with a standardized patient (actor) in an exam room. Your task will be to help ascertain which exams were performed.
<USER>
## EXAM STORY:
{exam_story}
## QUESTION:
During the exam described above with visual descriptions, is the physician performing an exam of the patient’s {exam_type}, yes or no? If so, describe what in the visual descriptions supports this answer in the fields below. If no, visual descriptions support, say “no visual evidence supports performance of exam”“.
## RESPONSE FORMAT:
Please ONLY provide a response in the following format as a JSON object with the “rationale” and “answer” keys and values as described.
{{
      “visual_evidence”: <summary of visual description evidence supporting presence or absence of exam>,
      “rationale”: <summary of rationale supporting final determination of whether the exam was performed>,
      “answer”: “yes” | “no”
~~~

4. Combined Input Physical Exam Assessment Prompt

~~~
<SYSTEM>
You are helpful teaching assistant at a medical school who is helping to assist in the grading of objective structured clinical examinations. You will be provided with summary descriptions of frames from a video recording of a medical student interacting with a standardized patient (actor) in an exam room. Your task will be to help ascertain which exams were performed.
<USER>
## EXAM STORY:
{exam_story}
## QUESTION:
During the exam described above, both with visual and audio descriptions, is the physician performing an exam of the patient’s {exam_type}, yes or no? If so, describe what in the visual descriptions or audio transcripts supports this answer in the fields below. If no, visual descriptions support, say “no visual evidence supports performance of exam”; if no audio transcripts support, say “no audio evidence supports performance of exam”.
## RESPONSE FORMAT:
Please provide a response in the following format as a JSON object with the “rationale” and “answer” keys and values as described.
{{
      “visual_evidence”: <summary of visual description evidence supporting presence or absence of exam>,
      “audio_evidence”: <summary of transcription evidence supporting presence or absence of exam>,
      “rationale”: <summary of rationale supporting final determination of whether the exam was performed>,
      “answer”: “yes” | “no”
}}
~~~

## D Multimodal Input Formatting Example

For the “Audio Only” exam story, each utterance is prefaced with its timestamp and concatenated in ascending timestamp order. For the “Visual Only”, each frame is prefaced with its timestamp followed by the extracted “student_posture”, “patient_posture”, “hand_movements”, and “tools” fields from the image descriptions and each frame description is concatenated in ascending timestamp order. For the “Combined” exam story, the relevant items are interleaved into one “story,” in ascending timestamp order, as illustrated in Figure 4 using the frame descriptions from LLaVA-1.6 34B.

**Figure 4:**
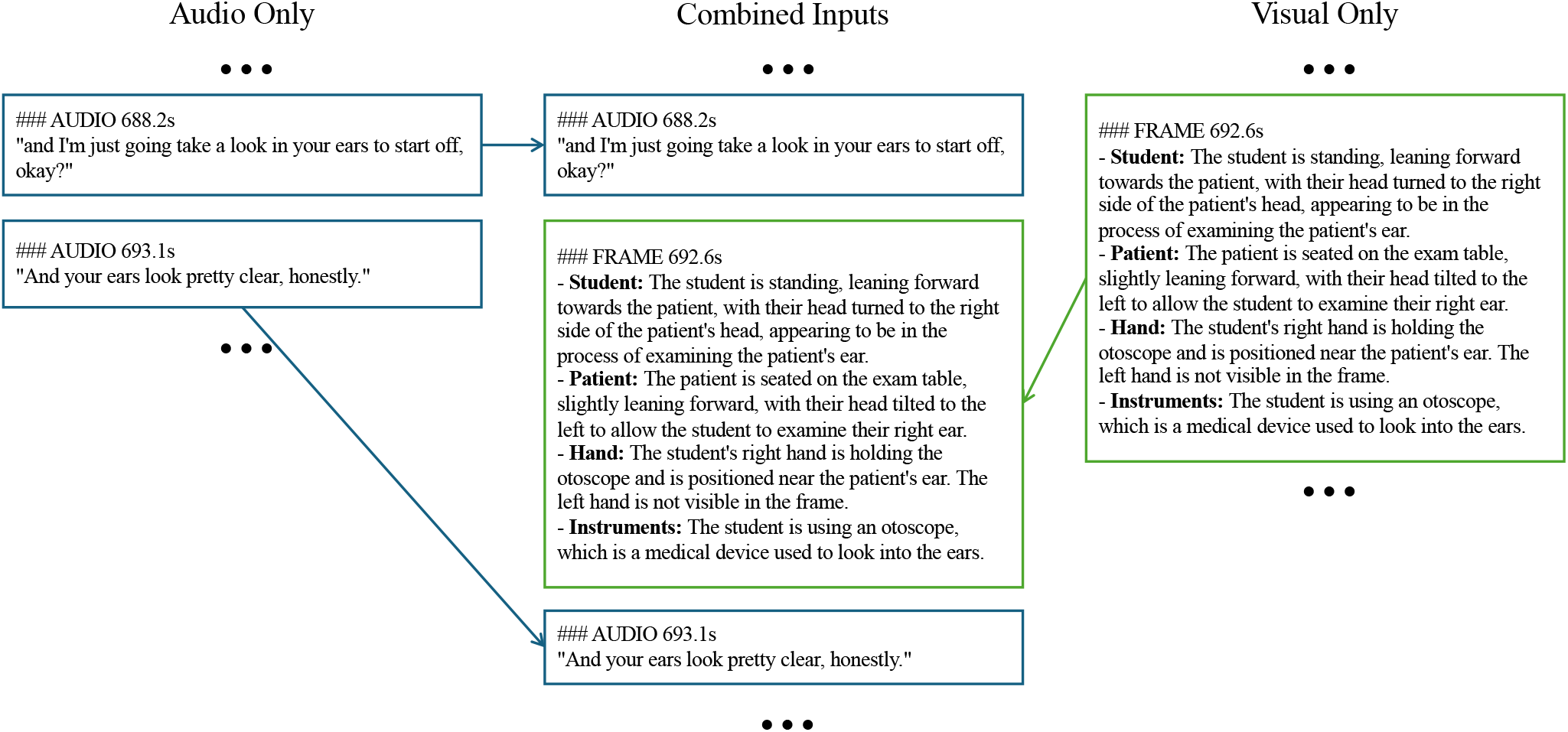
Example exam story excerpts

## E Example Responses

**Figure 5:**
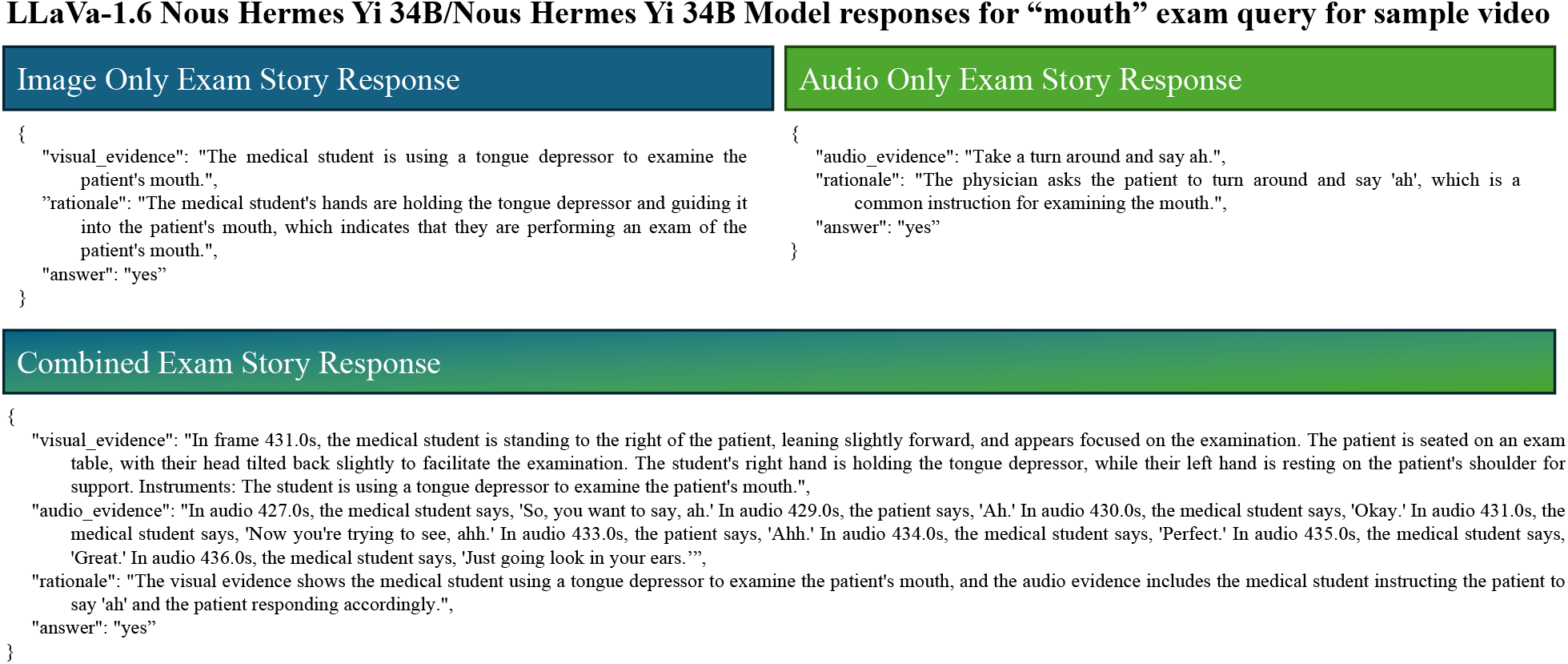
Sample Model Responses for Yi 34B for Mouth Exam

**Figure 6:**
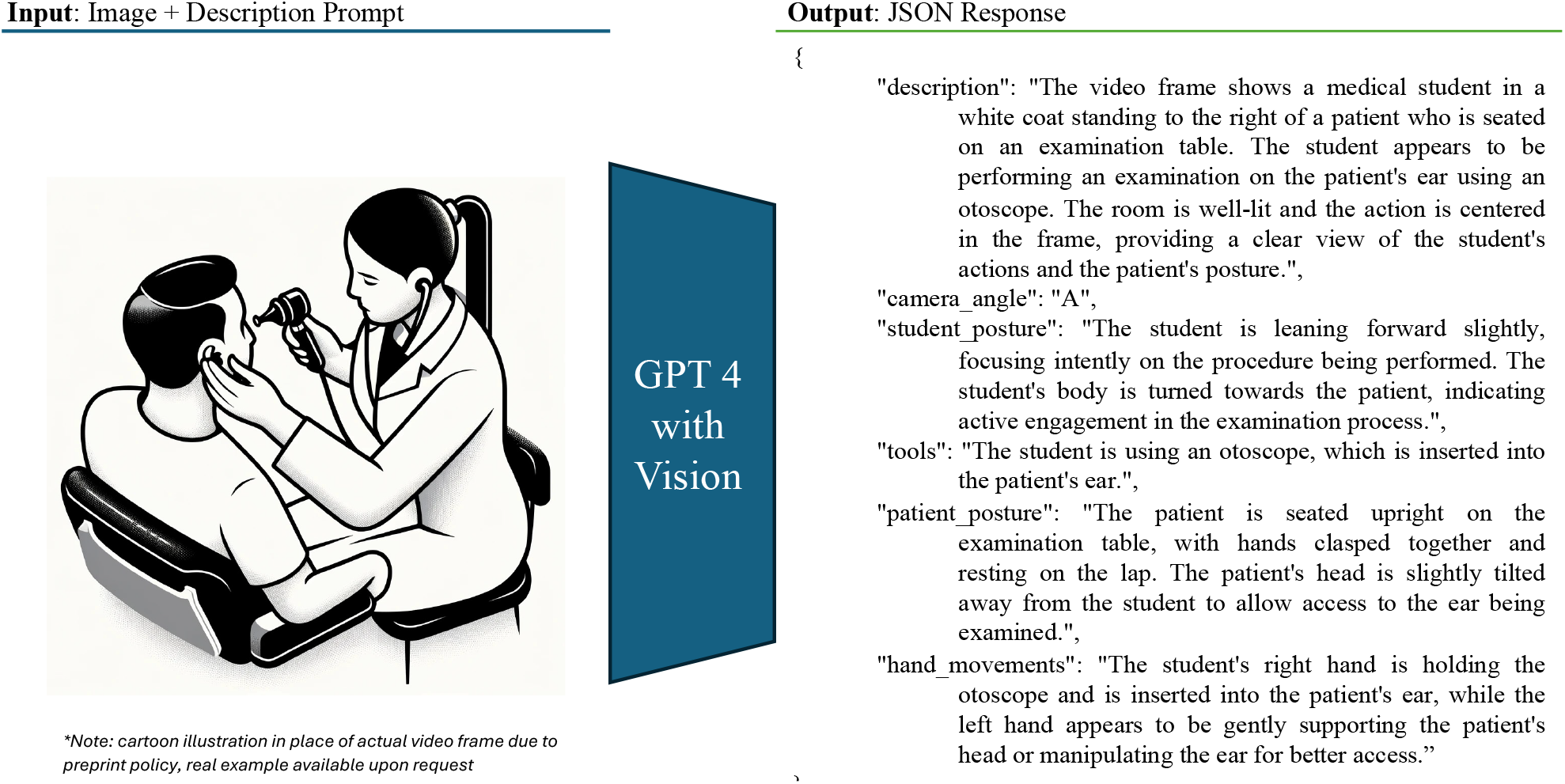
Example Output for Image Frame Description Prompt from GPT 4 with Vision

## F Image-Only Model Disagreement Mix

**Table 3:** SPE Disagreement Mix for Image-Only Model by Exam by Error Category.

| **Mouth Exam – Error Category** | **LLaVA**<br>**7B** | **LLaVA**<br>**13B** | **LLaVA**<br>**34B** | **GPT 4 w/**<br>**Vision** |
| --- | --- | --- | --- | --- |
| No tools, hands, or action detected | 73% | 79% | 51% | 82% |
| No tool and either no hands or no action | 15% | 19% | 46% | 11% |
| Tool or hands, but incorrect placement | 10% | 2% | 3% | 5% |
| Incorrect exam attributed | 1% | 0% | 0% | 1% |
| Incorrect tool detected | 0% | 0% | 0% | 1% |

| **Ear Exam – Error Category** | **LLaVA**<br>**7B** | **LLaVA**<br>**13B** | **LLaVA**<br>**34B** | **GPT 4 w/**<br>**Vision** |
| --- | --- | --- | --- | --- |
| No tools, hands, or action detected | 79% | 74% | 26% | 19% |
| No tool and either no hands or no action | 16% | 14% | 33% | 14% |
| Tool or hands, but no action detected | 4% | 5% | 7% | 9% |
| Incorrect exam attributed | 1% | 0% | 0% | 0% |
| Correct tool attributed to incorrect exam | 0% | 5% | 30% | 51% |
| Incorrect tool detected | 0% | 1% | 5% | 7% |

| **Nose Exam – Error Category** | **LLaVA**<br>**7B** | **LLaVA**<br>**13B** | **LLaVA**<br>**34B** | **GPT 4 w/**<br>**Vision** |
| --- | --- | --- | --- | --- |
| No tools, hands, or action detected | 81% | 81% | 49% | 83% |
| No tool and either no hands or no action | 6% | 13% | 49% | 9% |
| Tool or hands, but incorrect placement | 12% | 5% | 1% | 8% |
| Incorrect exam attributed | 1% | 1% | 1% | 0% |

## Notes

### Competing Interest Statement

The authors have declared no competing interest.

### Funding Statement

The authors greatly acknowledge Azure compute credits provided by Microsoft as part of a Accelerating Foundation Model Research grant award. (https://www.microsoft.com/en-us/research/project/afmr-cognition-and-societal-benefits/)

### Author Declarations

Ethics committee/IRB of UT Southwestern Medical Center gave ethical approval for this work.

### Summary of Updates

Added experimental results with GPT-4o.

## References

AI,:, Alex Young, Bei Chen, Chao Li, Chengen Huang, Ge Zhang, Guanwei Zhang, Heng Li, Jiangcheng Zhu, Jianqun Chen, Jing Chang, Kaidong Yu, Peng Liu, Qiang Liu, Shawn Yue, Senbin Yang, Shiming Yang, Tao Yu, et al. 2024. Yi: Open Foundation Models by 01.AI.

William F. Bond, Jianing Zhou, Suma Bhat, Yoon Soo Park, Rebecca A. Ebert-Allen, Rebecca L. Ruger, and Rachel Yudkowsky. 2023. Automated Patient Note Grading: Examining Scoring Reliability and Feasibility. Academic Medicine, 98(11S):S90–S97.

Wei-Lin Chiang, Zhuohan Li, Zi Lin, Ying Sheng, Zhanghao Wu, Hao Zhang, Lianmin Zheng, Siyuan Zhuang, Yonghao Zhuang, Joseph E. Gonzalez, Ion Stoica, and Eric P. Xing. 2023. Vicuna: An Open-Source Chatbot Impressing GPT-4 with 90%* ChatGPT Quality.

Tri Dao, Dan Fu, Stefano Ermon, Atri Rudra, and Christopher Ré. 2022. FlashAttention: Fast and Memory-Efficient Exact Attention with IO-Awareness. In S. Koyejo, S. Mohamed, A. Agarwal, D. Belgrave, K. Cho, and A. Oh, editors, Advances in Neural Information Processing Systems, volume 35, pages 16344–16359. Curran Associates, Inc.

Faiha Fareez, Tishya Parikh, Christopher Wavell, Saba Shahab, Meghan Chevalier, Scott Good, Isabella De Blasi, Rafik Rhouma, Christopher McMahon, Jean-Paul Lam, Thomas Lo, and Christopher W. Smith. 2022. A dataset of simulated patient-physician medical interviews with a focus on respiratory cases. Scientific Data, 9(1):313.

Albert Q. Jiang, Alexandre Sablayrolles, Arthur Mensch, Chris Bamford, Devendra Singh Chaplot, Diego de las Casas, Florian Bressand, Gianna Lengyel, Guillaume Lample, Lucile Saulnier, Lélio Renard Lavaud, Marie-Anne Lachaux, Pierre Stock, Teven Le Scao, Thibaut Lavril, Thomas Wang, Timothée Lacroix, and William El Sayed. 2023. Mistral 7B.

Woosuk Kwon, Zhuohan Li, Siyuan Zhuang, Ying Sheng, Lianmin Zheng, Cody Hao Yu, Joseph E. Gonzalez, Hao Zhang, and Ion Stoica. 2023. Efficient Memory Management for Large Language Model Serving with PagedAttention. In Proceedings of the ACM SIGOPS 29th Symposium on Operating Systems Principles.

Haotian Liu, Chunyuan Li, Qingyang Wu, and Yong Jae Lee. 2023. Visual Instruction Tuning.

Oyvind Meinich-Bache, Kjersti Engan, Ivar Austvoll, Trygve Eftestol, Helge Myklebust, Ladislaus Blacy Yarrot, Hussein Kidanto, and Hege Ersdal. 2020. Object Detection During Newborn Resuscitation Activities. IEEE Journal of Biomedical and Health Informatics, 24(3):796–803.

Meladel Mistica, Timothy Baldwin, Marisa Cordella, and Simon Musgrave. 2008. Applying discourse analysis and data mining methods to spoken OSCE assessments. In Proceedings of the 22nd International Conference on Computational Linguistics - COLING ‘08, volume 1, pages 577–584, Manchester, United Kingdom. Association for Computational Linguistics.

Nous Research. 2023. Nous-Hermes-2-Yi-34B. OpenAI, Josh Achiam, Steven Adler, Sandhini Agarwal, Lama Ahmad, Ilge Akkaya, Florencia

Leoni Aleman, Diogo Almeida, Janko Altenschmidt, Sam Altman, Shyamal Anadkat, Red Avila, Igor Babuschkin, Suchir Balaji, Valerie Balcom, Paul Baltescu, Haiming Bao, Mo Bavarian, Jeff Belgum, et al. 2023. GPT-4 Technical Report. 2303.08774 [cs].

Alec Radford, Jong Wook Kim, Tao Xu, Greg Brockman, Christine McLeavey, and Ilya Sutskever. 2023. Robust speech recognition via large-scale weak supervision. In International Conference on Machine Learning, pages 28492–28518. PMLR.

Jessica Salt, Polina Harik, and Michael A. Barone. 2019. Leveraging Natural Language Processing: Toward Computer-Assisted Scoring of Patient Notes in the USMLE Step 2 Clinical Skills Exam. Academic Medicine, 94(3):314–316.

Abeed Sarker, Ari Z. Klein, Janet Mee, Polina Harik, and Graciela Gonzalez-Hernandez. 2019. An interpretable natural language processing system for written medical examination assessment. Journal of Biomedical Informatics, 98:103268.

United States Medical Licensing Examination. n.d. Step 2 CK Content Outline & Specifications.

Sol Vedovato, Shinyoung Kang, Michael Holcomb, Krystle Campbell, Daniel Scott, Thomas Dalton, Gaudenz Danuser, and Andrew Jamieson. 2024. Towards better debriefing through context-aware video segmentation in standardized patient encounter ear exams. In pages 162–165, USA.

Thomas Wolf, Lysandre Debut, Victor Sanh, Julien Chaumond, Clement Delangue, Anthony Moi, Pierric Cistac, Tim Rault, Rémi Louf, Morgan Funtowicz, Joe Davison, Sam Shleifer, Patrick von Platen, Clara Ma, Yacine Jernite, Julien Plu, Canwen Xu, Teven Le Scao, Sylvain Gugger, et al. 2020. HuggingFace’s Transformers: State-of-the-art Natural Language Processing. 1910.03771 [cs].

Yuxin Wu, Alexander Kirillov, Francisco Massa, Wan-Yen Lo, and Ross Girshick. 2019. Detectron2.

Wen-wai Yim, Ashley Mills, Harold Chun, Teresa Hashiguchi, Justin Yew, and Bryan Lu. 2019. Automatic rubric-based content grading for clinical notes. In Proceedings of the Tenth International Workshop on Health Text Mining and Information Analysis (LOUHI 2019), pages 126–135, Hong Kong. Association for Computational Linguistics.

Lianmin Zheng, Wei-Lin Chiang, Ying Sheng, Siyuan Zhuang, Zhanghao Wu, Yonghao Zhuang, Zi Lin, Zhuohan Li, Dacheng Li, Eric P. Xing, Hao Zhang, Joseph E. Gonzalez, and Ion Stoica. 2023. Judging LLM-as-a-judge with MT-Bench and Chatbot Arena.

